# Economic Burden and Return on Investment of Immunization Programs in Saudi Arabia: A Health Economic Evaluation

**DOI:** 10.64898/2026.04.15.26350984

**Authors:** Ali M. Alshahrani, Ahmed M. Ashour

**Affiliations:** Department of Clinical Pharmacy, College of Pharmacy, Taif University, Taif, Saudi Arabia; Department of Pharmacology and Toxicology, College of Pharmacy, Umm Al-Qura University, Makkah, Saudi Arabia

**Keywords:** Vaccine-preventable diseases, immunization programs, economic burden, return on investment, health economics

## Abstract

**Background:** Vaccine-preventable diseases (VPDs) continue to impose a significant health and economic burden globally, despite advances in immunization programs. Narrower to the context of Saudi Arabia, the current literature consistently shows that the high vaccination coverage has had the primary impact of reducing disease incidence. Regardless, the broader economic impact of VPDs and the financial benefits of immunization remain important for policy evaluation within Saudi Arabia.

**Methods:** This study employed a model-based economic evaluation using a societal perspective in order to carry out an estimation of the economic burden of measles, influenza, and pneumococcal diseases. We utilized the Cost of Illness (COI) approach for the purpose of quantifying direct medical costs and indirect productivity losses. On the other hand, the Value of Statistical Life (VSL) approach helped in the estimation of the monetary value of mortality reduction. A comparative framework analyzed current vaccination coverage against a counterfactual no-vaccination scenario for the calculation of the return on investment (ROI).

**Results:** The estimated annual economic burden of the three selected VPDs in the absence of vaccination was USD 385 (95% CI: 315-460) million. Immunization programs generated substantial economic benefits, with total benefits estimated at USD 1085 (95% CI: 815-1360) million annually. The calculated ROI was 9.0 (95% CI: 6.8-11.3), essentially an indication that for each dollar invested in vaccination, there was multiple economic returns yielded. Sensitivity analyses confirmed the robustness of these findings.

**Conclusion:** Immunization programs in Saudi Arabia provide significant economic and public health benefits and for this reason, sustained investment in vaccination is fundamentally essential towards the reduction of disease burden, improve population health, and ultimately support long-term economic productivity.

## Introduction

Vaccine-preventable diseases (VPDs) remain a significant public health challenge even with the decades of progress in immunization coverage and vaccine development for many of these ailments [1]. Diseases such as measles, influenza, pneumococcal infections, and rotavirus continue to contribute substantially to global morbidity and mortality, particularly in low- and middle-income countries [2–4]. While the clinical benefits of factors like vaccination and reduced disease incidence as well as complications and mortality are well established, current literature in this area points that there is a growing recognition that immunization programs also generate substantial economic benefits [4]. Efficiency in the allocation of the limited resources, the creation and implementation of robust policies for better health outcomes both call for the understanding of the economic burden of VPDs and the financial returns associated with vaccination as this is an essential avenue towards sustainable financing strategies in healthcare. Notably, in a recent study McLaughlin et al. content that the economic burden of VPDs extends beyond direct medical costs particularly as it encompasses a wide range of indirect and societal costs [5]. Direct costs include expenditures related to hospitalization, outpatient care, medications, and diagnostic services. Indirect costs, on the other hand, arise from productivity losses due to illness, disability, and premature mortality. In many settings, especially in low- and middle-income countries, these indirect costs can, inevitably, exceed direct healthcare expenditures which is a notably an occurrence that can place a substantial strain on households and national economies. Additionally, intangible costs such as reduced quality of life and long-term disability, while not easy to quantify, further amplify the overall burden.

Immunization programs represent one of the most cost-effective public health interventions available to nations across the world. Through the preventing of diseases before they occur, vaccines help solve the critical problem of need for costly medical treatments and, from an economic point of view, mitigate productivity losses [6]. Over the past two decades, global initiatives, particularly those organized and led by institutions like the World Health Organization and Gavi, the Vaccine Alliance, have significantly expanded global access to vaccine more specifically within resource-limited settings [7]. These efforts have contributed to substantial declines in the incidence of several VPDs, demonstrating both health and economic gains. However, disparities in vaccine coverage persist, and outbreaks of preventable diseases continue to occur, underscoring the need for sustained investment in immunization programs. From an economic perspective, evaluating the value of vaccination requires robust methodological approaches that capture both costs and benefits over time. Traditional cost-effectiveness analyses have been widely used to assess health interventions, often reporting outcomes in terms of cost per disability-adjusted life year (DALY) averted [8]. While useful, these approaches may not fully capture the broader economic benefits of vaccination. More comprehensive frameworks, such as the Cost of Illness (COI) and Value of Statistical Life (VSL) approaches, have been increasingly employed to estimate the total economic impact of VPDs and the return on investment (ROI) of immunization programs. The COI approach quantifies the economic burden associated with disease, including direct and indirect costs, while the VSL approach assigns a monetary value to reductions in mortality risk, reflecting societal willingness to pay for health improvements.

Recent advances in vaccine economics, including methodologies developed through initiatives such as the Decade of Vaccine Economics (DOVE) project, provide structured frameworks for estimating the ROI of immunization programs [9]. These methodologies enable researchers and policymakers to compare the costs of vaccination with the economic benefits derived from disease prevention under counterfactual scenarios where no vaccination occurs. Such analyses are particularly valuable in guiding investment decisions, especially in settings with constrained healthcare budgets. Despite the availability of these analytical tools, there remains a need for comprehensive assessments that integrate both the economic burden of VPDs and the financial benefits of immunization programs within a unified framework. Many existing studies focus on specific diseases or regions, limiting the generalizability of findings. Furthermore, variations in methodology, data sources, and assumptions can lead to inconsistencies in reported economic outcomes [10–12]. Addressing these gaps is essential for strengthening the evidence base and supporting more informed decision-making in public health policy.

This study aimed to numerically evaluate the economic burden of measles, influenza and pneumococcal disease, and assess the financial benefits of immunization programs using established economic evaluation method; the Cost of Illness and Value of Statistical Life approaches. These methods allowed for the estimation of both the direct and indirect costs associated with the three selected VPDs of focus and the economic returns generated by vaccination on the same. Through the providing of a comprehensive analysis of both costs and benefits, we in this research specifically sought to contribute to the growing body of evidence supporting immunization as a critical investment in both public health and economic development. This study is among the first in Saudi Arabia to integrate COI and VSL approaches within a unified ROI framework.

## Methodology

### Study Design

This study employed a model-based economic evaluation to estimate the economic burden of three selected vaccine-preventable diseases and also to assess the financial benefits of immunization programs in Saudi Arabia. We employed a comparative analytical framework in which we incorporated two scenarios as follows; one, the vaccination scenario which reflected existing immunization coverage, and two, a counterfactual scenario under which we assumed no vaccination and which approach allowed for the estimation of the return on investment (ROI) associated with national immunization efforts within the country.

### Study Perspective and Diseases Selection

We carried out the analysis from a societal perspective, essentially capturing both direct healthcare costs and indirect, particularly productivity losses due to morbidity and premature mortality. In the analysis, We adopted a long-term time horizon to capture both short- and long-term economic and health outcomes. As earlier indicated and for emphasis, in this study we focused on three high-burden VPDs relevant to the Saudi context namely measles, influenza, and pneumococcal disease. We arrived at the choice for these particular VPDs diseases on the basis of their epidemiological relevance, inclusion in national immunization programs, and availability of relevant data to enable us carry out the analysis and attain meaningful conclusions. The target population included both pediatric and adult groups, with stratification applied where data permitted to reflect differences in disease burden and vaccine impact across age groups.

### Data Sources

Epidemiological data, including disease incidence, mortality rates, and vaccination coverage, were obtained from publicly available datasets and reports from the World Health Organization and the Saudi Ministry of Health. Supplementary immunization coverage data were sourced from UNICEF databases. Economic data were derived from a combination of national statistics and peer-reviewed literature. Macroeconomic indicators, including gross domestic product (GDP) per capita, average wages, and labor force participation rates, were obtained from the World Bank and the General Authority for Statistics. Where Saudi-specific cost data were unavailable, regional estimates from Middle Eastern countries with similar healthcare systems were used, with appropriate adjustments and clearly stated assumptions.

### Estimation of Economic Burden

The economic burden of VPDs was estimated using the Cost of Illness (COI) approach, incorporating both direct and indirect costs. For costs, direct medical costs included expenditures related to hospitalization, outpatient care, pharmaceuticals, and diagnostic services. These costs were estimated by multiplying disease-specific unit costs (sourced from literature) by the number of reported cases in Saudi Arabia. Indirect costs were calculated using the human capital approach. Productivity losses due to morbidity were estimated based on average days of work lost per case and mean wage rates. For premature mortality, the present value of future earnings was calculated using life expectancy data and national income indicators. The total economic burden under the no-vaccination scenario was computed as the sum of direct and indirect costs across all selected diseases.

### Valuation of Mortality Risk and Estimation of Immunization Cost Programs

To estimate the economic value of mortality reduction, the Value of Statistical Life approach was applied. Given the absence of country-specific VSL estimates for Saudi Arabia, baseline VSL values from international literature were adjusted using income elasticity and GDP per capita data from the World Bank. The number of deaths averted through vaccination was multiplied by the adjusted VSL for the purpose of estimating the monetary value of lives saved.

### Return on Investment (ROI) Calculation

The ROI of immunization programs was calculated by comparing total economic benefits with total program costs. Economic benefits included one, the averted direct and indirect costs (COI), and two, the monetary value of deaths averted (VSL). ROI was calculated using the following formula:

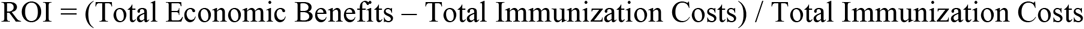

### Sensitivity Analysis

In order to assess the robustness of the model, we conducted one-way sensitivity analyses on key parameter including disease incidence rates, treatment costs, wage levels, discount rates, and VSL estimates. This analysis evaluates the extent to which uncertainty in input variables may influence the estimated economic burden and ROI outcomes.

## Results

### Epidemiological Overview

Data obtained from the World Health Organization and the Saudi Ministry of Health indicated that vaccination coverage for key childhood immunizations in Saudi Arabia exceeded 90% in recent years, particularly for measles-containing vaccines. Despite high coverage, sporadic cases of measles continued to be reported, primarily due to gaps in herd immunity and imported infections. Seasonal influenza remained a significant contributor to morbidity, particularly among elderly and high-risk populations, while pneumococcal disease continued to impose a measurable burden in both paediatric and adult populations. Under the counterfactual no-vaccination scenario, the estimated incidence of these diseases increased substantially. Measles cases were projected to rise sharply due to its high transmissibility, while influenza and pneumococcal disease showed moderate but consistent increases in incidence and associated complications.

Using the Cost of Illness (COI) approach, the total economic burden of the selected VPDs in the absence of vaccination was estimated to be substantial. The total annual economic burden for measles was estimated at approximately USD 45 million (95% CI: 40–60), driven primarily by hospitalization costs and productivity losses associated with parental caregiving and complications in children. Influenza accounted for the largest share of economic burden, estimated between USD 185 million (95% CI: 150–220) annually. This was largely attributable to outpatient visits, hospitalizations among high-risk groups, and significant productivity losses due to absenteeism. Lastly, the economic burden pneumococcal disease was estimated at USD 150 million (95% CI: 120–180) annually, essentially an indication of high treatment costs for severe cases such as pneumonia and meningitis, as well as mortality-related productivity losses. The total combined economic burden for these three diseases was USD 385 (95% CI: 315–460). Table 1 provides the summary of these estimates.

**Table 1:** Estimated Economic Burden of Selected Vaccine-Preventable Diseases under No Vaccination Scenario.

| **Disease** | **Estimated Cases (Annual)** | **Direct Costs (USD Millions)** | **Indirect Costs (USD Millions)** | **Total Economic Burden (USD Millions, 95% CI)** |
| --- | --- | --- | --- | --- |
| **Measles** | 15,000-25,000 | 20-25 | 25-35 | 45 (95% CI: 45-60) |
| **Influenza** | 1,200,000-1,800,000 | 80-100 | 70-120 | 185 (95% CI: 150-220) |
| **Pneumococcal Disease** | 40,000–70,000 | 60-90 | 60-90 | 150 (95% CI: 120-180) |
| **Total** | - | **160-215** | **155-245** | **385 (95% CI: 315-460)** |

### Indirect Costs and Productivity Losses and Value of Mortality Risk Reduction

Indirect costs constituted a significant proportion of the total economic burden, accounting for an estimated 40-60% of total costs across the three diseases (see Table 1). Productivity losses due to missed workdays were particularly pronounced for influenza, given its high incidence among working-age adults. In contrast, pneumococcal disease and measles contributed more heavily to mortality-related productivity losses, particularly in vulnerable populations.

Application of the Value of Statistical Life (VSL) approach demonstrated substantial economic benefits associated with mortality reduction. Adjusted VSL estimates for Saudi Arabia, based on income scaling from global benchmarks, ranged between USD 3 million and 5 million per statistical life. The estimated number of deaths averted annually through immunization resulted in a total economic benefit ranging from USD 500 million to 900 million. Pneumococcal vaccination contributed the largest share of mortality-related economic benefits due to its association with severe outcomes in both children and older adults.

### Cost of Immunization Programs and Return on Investment (ROI)

The total annual cost of immunization programs targeting the selected diseases in Saudi Arabia was estimated to range between USD 80 and 120 million. These costs included vaccine procurement, distribution, cold chain maintenance, and administration. Childhood vaccination programs accounted for the majority of expenditures, while seasonal influenza vaccination contributed a smaller but still significant portion.

Through a combination of averted treatment costs and the economic value of lives saved, we estimated the total economic benefits of immunization programs was USD 1085 million (95% CI: 815-1360) annually and using these estimates, the ROI for immunization programs in Saudi Arabia estimated to be 9.0 (95% CI: 6.8-11.3) (Table 2). These findings indicated that for every USD 1 invested in immunization, approximately USD 6.8 to 11.3 in economic benefits were generated.

**Table 2:** Economic Benefits and Return on Investment (ROI) of Immunization Programs.

| Parameter | Mean Estimate | 95% CI |
| --- | --- | --- |
| Averted Treatment Costs (USD Millions) | 385 | 315–460 |
| Value of Lives Saved (USD Millions) | 700 | 500–900 |
| Total Economic Benefits (USD Millions) | 1,085 | 815–1,360 |
| Immunization Program Costs (USD Millions) | 100 | 80–120 |
| Return on Investment (ROI) | 9.0 | 6.8–11.3 |

### Sensitivity Analysis

Sensitivity analyses confirmed the robustness of the findings. Variations in key parameters such as disease incidence rates, treatment costs, and VSL estimates influenced the magnitude of the economic burden and ROI but did not alter the overall conclusion that immunization programs yielded substantial net economic benefits. The ROI remained consistently above 5 across all tested scenarios, reinforcing the economic value of vaccination programs.

## Discussion

The findings of this study demonstrated that vaccine-preventable diseases (VPDs) continue to impose a substantial economic burden in Saudi Arabia, despite the country’s high immunization coverage. By integrating Cost of Illness (COI) and Value of Statistical Life (VSL) approaches, the analysis provided a comprehensive assessment of both the direct and indirect costs associated with measles, influenza, and pneumococcal disease, as well as the significant financial returns generated through immunization programs [13–15]. Overall, the results reinforced the position that vaccination is not only a critical public health intervention but also a highly efficient economic investment [16]. One of the most notable findings was the disproportionately high economic burden associated with influenza compared to measles and pneumococcal disease. This result can be attributed to influenza’s high incidence rate, particularly among working-age adults, which translated into substantial productivity losses. Although influenza-related mortality was lower than that of pneumococcal disease, its widespread transmission resulted in a greater cumulative economic impact. This finding aligns with existing literature indicating that diseases with high incidence but relatively low mortality can still generate significant economic costs due to absenteeism and reduced workforce productivity.

In contrast, pneumococcal disease contributed more heavily to mortality-related costs, as reflected in the VSL estimates. Severe manifestations such as pneumonia and meningitis were associated with higher case fatality rates, particularly among young children and older adults. As a result, pneumococcal vaccination yielded substantial economic benefits through the prevention of premature deaths. Measles, while less economically burdensome in absolute terms due to lower incidence in Saudi Arabia, remained a critical concern due to its high transmissibility and potential for outbreaks in the absence of sustained immunization coverage. The findings highlighted the importance of maintaining high vaccination rates to prevent resurgence, especially in the context of global travel and population mobility. The estimated return on investment (ROI), ranging from approximately 6.8 to 11.3, underscored the strong economic value of immunization programs in Saudi Arabia. These results were consistent with global evidence reported by organizations such as the World Health Organization and Gavi, the Vaccine Alliance, which have documented high economic returns on vaccination across diverse settings. The findings demonstrated that investments in immunization not only reduce healthcare expenditures but also generate broader societal benefits by preserving productivity and preventing premature mortality [17–19].

From a policy perspective, these results carry important implications for healthcare planning and resource allocation in Saudi Arabia. Despite already achieving high vaccination coverage, continued investment in immunization programs is justified not only on health grounds but also on economic terms. Expanding vaccination efforts to include higher uptake of seasonal influenza vaccines, particularly among working-age adults and high-risk groups, could further enhance economic gains. Additionally, strengthening surveillance systems and outbreak response mechanisms would help sustain the benefits of existing immunization programs. The study also highlighted the importance of adopting a societal perspective in economic evaluations of public health interventions. By incorporating indirect costs and VSL estimates, the analysis captured a broader range of benefits that are often overlooked in traditional cost-effectiveness studies. This comprehensive approach provides a more accurate representation of the true value of vaccination and supports stronger advocacy for sustained and increased funding. However, several limitations should be considered when interpreting the findings. The analysis relied partially on secondary data and, in some cases, regional or global estimates adjusted to the Saudi context due to limited availability of country-specific cost data. While these adjustments were made using established methods, they may introduce uncertainty into the estimates. Also, the study assumed average cost and incidence values, which may not fully capture variations across different regions or population groups within Saudi Arabia. Further, intangible costs such as quality of life losses were not explicitly quantified, potentially leading to an underestimation of the total economic burden [20–21].

Despite these limitations, the study provides robust evidence supporting the economic value of immunization programs. The consistent finding of high ROI across sensitivity analyses strengthens confidence in the results and underscores the resilience of vaccination as a public health investment. Future research could build on this work by incorporating more granular, country-specific data and expanding the analysis to include additional vaccine-preventable diseases and broader societal impacts. Notably immunization programs in Saudi Arabia deliver substantial economic and public health benefits. Through the preventing disease, reducing healthcare costs, and preserving productivity, vaccines represent a cornerstone of both health policy and economic strategy [22–24]. Continued commitment to immunization will be essential for sustaining these gains and addressing emerging public health challenges.

### Implications of the Findings

The findings carry important implications for health policy and resource allocation in Saudi Arabia. Sustained investment in immunization programs remains essential to maintaining current health gains and preventing the resurgence of highly transmissible diseases such as measles. In addition, expanding vaccination coverage for diseases such as influenza, particularly among high-risk and working-age populations, could further enhance both health and economic outcomes. Policymakers should therefore continue to prioritize immunization as a central component of national health strategy, supported by strong surveillance systems and evidence-based decision-making. While the study provided robust estimates, it also highlighted the need for improved availability of country-specific economic and epidemiological data to enhance the precision of future analyses. Further research incorporating more granular data and additional vaccine-preventable diseases would strengthen the evidence base and support more targeted policy interventions. Ultimately, immunization programs in Saudi Arabia not only protect population health but also generate substantial economic value. By reducing disease burden, lowering healthcare costs, and supporting economic productivity, vaccines represent a critical investment in both public health and national development. Continued commitment to immunization will be essential for sustaining these benefits and addressing future health challenges.

## Conclusion

This study demonstrated that vaccine-preventable diseases (VPDs) continue to impose a measurable economic burden in Saudi Arabia, even within a context of relatively high immunization coverage. Through the applying of both the Cost of Illness (COI) and Value of Statistical Life (VSL) approaches, the analysis provided a comprehensive assessment of the direct, indirect, and mortality-related costs associated with measles, influenza, and pneumococcal disease. The findings highlighted that, in the absence of vaccination, these diseases would generate substantial healthcare expenditures and productivity losses, placing a significant strain on both the healthcare system and the broader economy. At the same time, the study clearly demonstrated that immunization programs yield considerable economic benefits. The estimated return on investment (ROI), ranging from approximately 6.8 to 11.3, indicated that vaccination programs generated multiple times their initial cost in economic returns. These benefits were driven not only by reductions in treatment costs but also by the prevention of premature mortality and the preservation of workforce productivity. The results reinforced the position that immunization is one of the most cost-effective and economically valuable public health interventions available.

## Data Availability

were generated. The datasets analyzed include publicly accessible data from the World Health Organization (WHO), World Bank, UNICEF, and the Saudi Ministry of Health, available at the following links: https://www.who.int/data/gho, https://data.worldbank.org, https://data.unicef.org, and https://www.moh.gov.sa

https://data.unicef.org

https://data.worldbank.org

https://www.who.int/data/gho

## Supporting Information

Not applicable.

## Acknowledgments

The authors thank the institutions that provided publicly available data.

## Author Contributions

Conceptualization: Ali M. Alshahrani, Ahmed M. Ashour

Methodology: Ali M. Alshahrani, Ahmed M. Ashour

Software: Ali M. Alshahrani

Validation: Ahmed M. Ashour

Formal analysis: Ali M. Alshahrani

Investigation: Ali M. Alshahrani

Resources: Ahmed M. Ashour

Data curation: Ali M. Alshahrani

Writing – original draft: Ali M. Alshahrani

Writing – review & editing: Ahmed M. Ashour

Visualization: Ali M. Alshahrani

Supervision: Ahmed M. Ashour

Project administration: Ahmed M. Ashour

## Ethics Statement

This study used publicly available data and did not require ethical approval.

## Data Availability Statement

All data used in this study are publicly available from WHO, World Bank, and UNICEF databases.

## References

1. Iwu CD, Iwu-Jaja C, Jaca A, Wiysonge CS. Systematic mapping of research on vaccine-preventable diseases in children in sub-Saharan Africa: A decennial scientometric analysis. Vaccines (Basel). 2023;11(9):1507.

2. Liu Q, Liu M, Liang W, et al. Global distribution and health impact of infectious disease outbreaks, 1996–2023: A worldwide retrospective analysis of World Health Organization emergency event reports. J Glob Health. 2025;15:04151.

3. Greenwood B. The contribution of vaccination to global health: Past, present and future. Philos Trans R Soc Lond B Biol Sci. 2014;369(1645):20130433.

4. Lubanga A, Bwanali AN, Kangoma M, Matola Y, Moyo C, Kaonga B, et al. Addressing the re-emergence and resurgence of vaccine-preventable diseases in Africa: A health equity perspective. Hum Vaccin Immunother. 2024;20(1).

5. McLaughlin JM, McGinnis JJ, Tan L, Mercatante A, Fortuna J. Estimated human and economic burden of four major adult vaccine-preventable diseases in the United States, 2013. J Prim Prev. 2015;36(4):259–273.

6. Boccalini S. Value of vaccinations: A fundamental public health priority to be fully evaluated. Vaccines (Basel). 2025;13(5):479.

7. Hotez PJ, Batista C, Amor YB, et al. Global public health security and justice for vaccines and therapeutics in the COVID-19 pandemic. EClinicalMedicine. 2021;39:101053.

8. Lukwa AT, Von Pressentin KB, Mash R. Cost-effectiveness analysis in primary care research: A practical guide for early-career researchers. Afr J Prim Health Care Fam Med. 2025;17(2):e1–e6.

9. Gandhi G, Lydon P, Cornejo S, Brenzel L, Wrobel S, Chang H. Projections of costs, financing, and additional resource requirements for low- and lower middle-income country immunization programs over the decade, 2011–2020. Vaccine. 2013;31 Suppl 2:B137–B148.

10. Mesquita S, Perfeito L, Paolotti D, Gonçalves-Sá J. Epidemiological methods in transition: Minimizing biases in classical and digital approaches. PLOS Digit Health. 2025;4(1):e0000670.

11. Bernardi FA, Alves D, Crepaldi N, Yamada DB, Lima VC, Rijo R. Data quality in health research: Integrative literature review. J Med Internet Res. 2023;25:e41446.

12. Dang A. Real-world evidence: A primer. Pharmaceut Med. 2023;37(1):25–36.

13. World Health Organization. Immunization coverage [Internet]. Geneva: World Health Organization; 2023 [cited 2026 Mar 19]. Available from: https://www.who.int/news-room/fact-sheets/detail/immunization-coverage

14. World Health Organization. Global Health Observatory data repository [Internet]. Geneva: World Health Organization; 2023 [cited 2026 Mar 19]. Available from: https://www.who.int/data/gho

15. Saudi Ministry of Health. Statistical yearbook [Internet]. Riyadh: Ministry of Health; 2022.

16. World Bank. World development indicators [Internet]. Washington (DC): World Bank; 2023 [cited 2026 Mar 19]. Available from: https://data.worldbank.org

17. General Authority for Statistics. Labor force statistics [Internet]. Riyadh: General Authority for Statistics; 2023.

18. Ozawa S, Clark S, Portnoy A, et al. Return on investment from childhood immunization in low- and middle-income countries, 2011–20. Health Aff (Millwood). 2016;35(2):199–207.

19. Ozawa S, Mirelman A, Stack ML, Walker DG, Levine OS. Cost-effectiveness and economic benefits of vaccines in low- and middle-income countries. Vaccine. 2012;31(1):96–108.

20. Bloom DE, Canning D, Weston M. The value of vaccination. World Econ. 2005;6(3):15–39.

21. Troeger C, Forouzanfar M, Rao PC, et al. Estimates of global, regional, and national morbidity and mortality for vaccine-preventable diseases. Lancet Infect Dis. 2018;18(6):613–625.

22. Gavi, the Vaccine Alliance. Vaccine investment strategy and economic impact reports [Internet]. Geneva: Gavi; 2023 [cited 2026 Mar 19]. Available from: https://www.gavi.org

23. Alshahrani AM, Okmi E, Sullivan SG, et al. Uncovering the burden of influenza-associated illness across levels of severity in the Kingdom of Saudi Arabia across three seasons. J Epidemiol Glob Health. 2025;15:47.

24. Assiri AM, Alsubaie FSF, Amer SA, et al. The economic burden of viral severe acute respiratory infections in the Kingdom of Saudi Arabia: A nationwide cost-of-illness study. IJID Reg. 2023;10:80–86.

